# US Overdose Mortality Saw First Drop Below the Jalal-Burke Exponential Growth Curve in 2024

**DOI:** 10.1101/2025.10.24.25338732

**Authors:** Joseph R. Friedman, Joseph J. Palamar, Daniel Ciccarone, Tommi L. Gaines, Annick Borquez, Chelsea L. Shover, Steffanie A. Strathdee

**Author notes:** Correspondence to: Joseph Friedman, MD, PhD, MPH, University of California, San Diego, 200 W Arbor Dr, San Diego, CA, 92103.

## Abstract

**Background:** Between 1979 and 2016, US overdose death rates rose in a smooth fashion, described by Jalal and Burke using an exponential growth curve that fit observed data nearly perfectly. Fluctuations above this curve have subsequently been seen during shocks related to drug supply and the COVID-19 pandemic. However, large-magnitude dips below the curve have never been demonstrated. Given that overdose mortality began sharply falling during 2023-2024, we assess updated overdose trends against the Jalal-Burke curve.

**Methods:** We examined US overdose deaths from the National Vital Statistics System between January 1979-December 2024. We recreated the Jalal-Burke curve, fitting an exponential growth curve to overdose rates from 1979 to 2016, linearly projecting through 2024, with 95% confidence intervals. We also examined trends by specific substance involvement.

**Results:** After precipitously surpassing exponential growth predictions in 2020-2023, overdose deaths decreased sharply from approximately 32 per 100,000 in 2021-2023 to 23.7 in 2024, falling below the lower bound of Jalal-Burke curve (24.98 per 100,000) for the first time since 2001. These decreases reflected declining illicit fentanyl-involved deaths (with and without stimulants); however, deaths involving stimulants without fentanyl, and those involving xylazine, represent an increasing share of deaths in 2024.

**Conclusions:** Rather than simply representing a return to the Jalal-Burke exponential growth curve, recent decreases in overdose deaths represent the first significant, large-magnitude deviation below exponential growth projections. These trends represent a very positive development. However, challenges in the US drug crisis are shifting, requiring a tailored public health response.

## Introduction

Drug overdose mortality in the United States (US) has risen steadily for decades. In their landmark study in *Science*, Jalel et al. described this phenomenon, and showed that the US overdose death rate between 1979 and 2016 could be described in a nearly mathematically perfect fashion with a simple exponential growth curve (reflecting an R^2^ value of 0.99) (Jalal et al., 2018). This insight about the nature of the overdose crisis sparked scientific debate about the mechanisms underpinning exponential growth, and the durability of similar trends into the future (Caulkins, 2022; Compton et al., 2022; Keyes & Cerdá, 2022; Reuter, 2022).

In subsequent analyses incorporating additional years of data, Jalal and Burke—as well as other author groups—have highlighted that deviations above this curve have occurred in the context of drug supply shocks and the COVID-19 pandemic (Compton et al., 2022; Jalal & Burke, 2022, 2026). Nevertheless, large-magnitude shifts below the Jalal-Burke curve have never been demonstrated. However, given that the US overdose death rate began decreasing in mid-2023 (J. R. Friedman et al., 2026; Post et al., 2025), we assess updated overdose mortality trends against the Jalal-Burke curve.

An additional key insight from Jalal et al. was that, despite the smooth overall growth of the US overdose mortality curve, it was composed of a shifting profile of drugs over time (Jalal et al., 2018). This shifting composition, over the past 25 years, has also been described using a four-wave paradigm (Ciccarone, 2021). Waves 1-4 correspond roughly to deaths from prescription opioids, heroin, synthetic opioids (mainly illicitly manufactured fentanyl) without stimulants (including psychostimulants, e.g., methamphetamine, and cocaine), and fentanyl with stimulants, respectively (Ciccarone, 2021; J. R. Friedman et al., 2026; J. Friedman & Shover, 2023). Here, we leverage substance-specific overdose mortality data to examine how each of these waves has contributed to overall progress against the Jalal-Burke curve.

## Methods

Nationally representative drug overdose death data were obtained from the National Vital Statistics System for 1979-2024. Overdose mortality was defined using ICD-10 underlying cause of death codes X40–X44 (unintentional), X60–X64 (suicide), X85 (homicide), and Y10–Y14 (undetermined). Of note, the original analysis by Jalal et al. used overdose mortality data corresponding only to accidental overdoses (X40—X44) (Jalal et al., 2018), which do represent the vast majority of overdose deaths, but misses those corresponding to suicide, homicide, or unknown intent. For instance, in 2024, accidental overdoses represented about 92% of total observed overdose deaths (*CDC WONDER*, n.d.). Follow-up studies (Compton et al., 2022), including most federal studies and databases (Ahmad et al., 2020), and follow-up studies by Jalal and Burke (Jalal & Burke, 2026) have tended to include the broader set of codes, which we also employed here for consistency with the most recent literature. This is not expected to have a large impact on the direction of findings or magnitude of trends compared to analyzing only accidental overdoses.

Data on deaths between 1999 and 2024 were obtained for overdose-related deaths overall and stratified by year and substance-involvement via the Centers for Disease Control and Prevention (CDC) Wide-ranging ONline Data for Epidemiologic Research (WONDER) platform (CDC WONDER, n.d.). As the CDC WONDER platform does not provide records before 1999, mortality rates at the national level between 1979 and 1998 were obtained by year from a separate publication using NVSS data (Compton et al., 2022).

Trends broken down by substance involvement were obtained for prescription opioids (T40.2), heroin (T40.1), synthetic opioids (primarily and herein referred to as fentanyl) [T40.4], cocaine (T40.5), psychostimulants with abuse potential (primarily and herein referred to as methamphetamine) [T43.6], and xylazine (T42.7 or T46.5).

To recreate the Jalal-Burke exponential curve, we fit a bivariate linear regression model with the outcome as the natural log of deaths per 100,000 population and the explanatory variable of year of occurrence, using national overdose death data from 1979 to 2016. This represents the same time period as the original analysis (Jalal et al., 2018). The model used to fit data between 1979 and 2016 was linearly extrapolated to make predictions for the 2017-2024 period. Observed trends between 1979 and 2024 were compared with model predictions, with 95% confidence intervals (CIs), drawn from the fitted model assuming a normal distribution.

Analyses were conducted in R version 4.4.1. This analysis of deidentified data was exempt from review by the University of California, San Diego, Institutional Review Board.

## Results

During the COVID-19 pandemic in 2020, overdose mortality rose sharply above the exponential growth curve (Figure 1). Rates remained elevated above the curve prediction, at approximately 32 deaths per 100,000 population between 2021 and 2023. In 2024, national overdose mortality fell precipitously to 23.7 per 100,000, dropping below the 95% CI of the exponential curve prediction (24.1-29.0 per 100,000) for the first time since 2001—and for the first time ever to a large magnitude degree.

**Figure 1.**
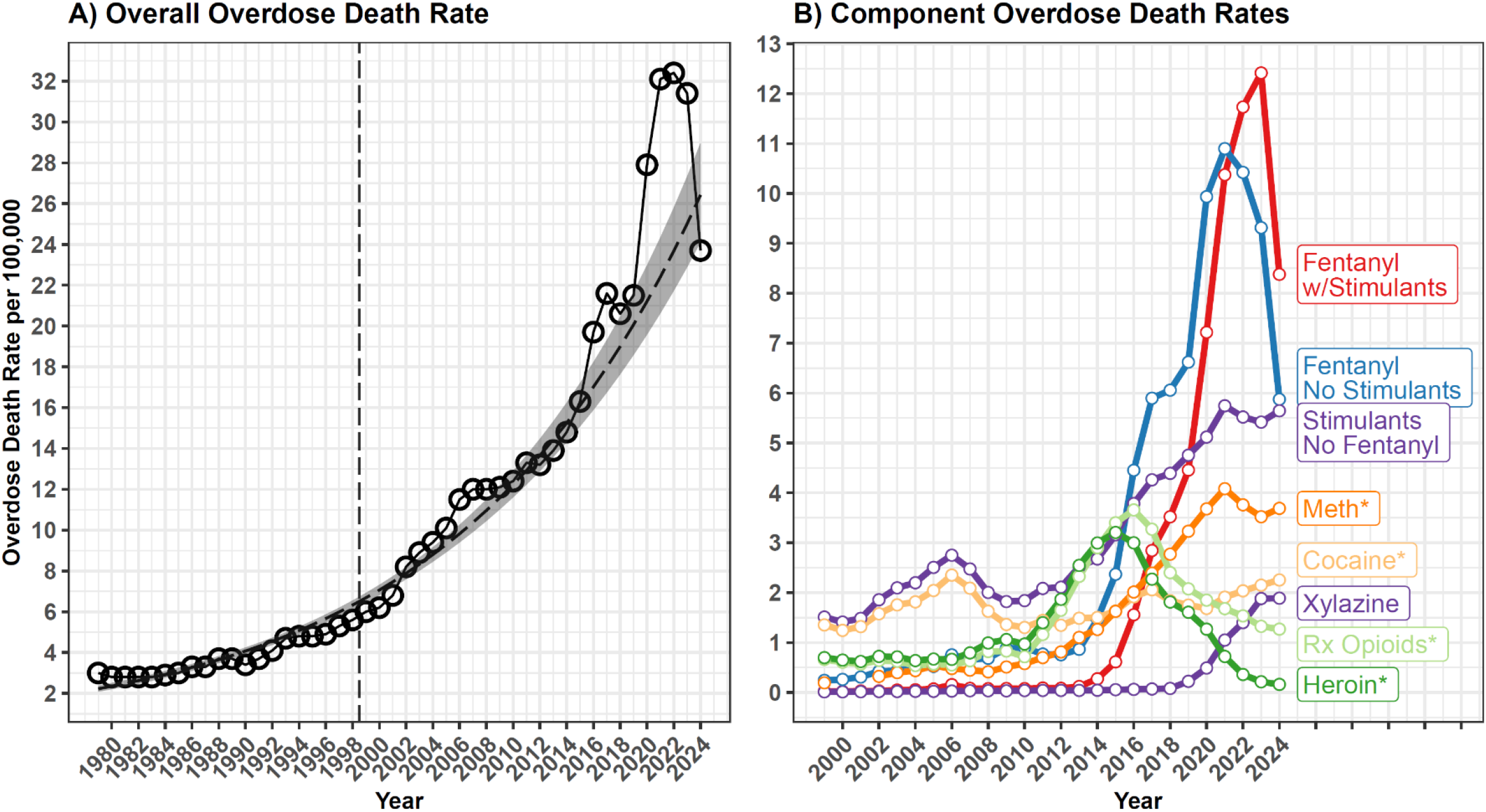
Overdose Death Rates Compared to Exponential Growth and by Substance Co-Involvement. A) National-level estimates of overdose deaths per 100,000 are shown by year from 1979 to 2024. Following the general approach established by Jalal et al., an exponential line of best fit is shown, estimated using data from 1979-2016 with a 95% uncertainty interval. The vertical dashed line shows the portion of the graph corresponding to figure part B, for which drug-specific data were available from 1999-2024. B) Drug overdose death rates per 100,000 are shown by specific substance involvement. Fentanyl-involved deaths are shown separately by stimulant co-involvement status. *Deaths for methamphetamine (meth), cocaine, commonly prescribed opioids (Rx Opioids) and heroin are shown removing deaths co-involved with fentanyl. Data from 2024 are provisional and many slightly underestimate final numbers.

Rates of overdose deaths involving prescription opioids (with fentanyl co-involved deaths omitted), heroin (fentanyl omitted), fentanyl (without stimulants), and fentanyl (with stimulants) reached inflection points and began to fall in 2017, 2016, 2022, and 2024, respectively, marking the decline of waves 1-4 of the crisis, respectively. Cocaine, methamphetamine, and xylazine reflect segments of the overdose curve that were not declining in 2024. In 2024 the rate of deaths involving stimulants without fentanyl (5.64 per 100,000) almost caught up to the rate of deaths involving fentanyl without stimulants (5.87 per 100,000). This reflects that deaths involving methamphetamine (without fentanyl) increased from 3.52 per 100,000 in 2023 to 3.69 per 100,000 in 2024, and deaths involving cocaine (without fentanyl) increased from 2.14 per 100,000 in 2023 to 2.25 per 100,000 in 2024. Deaths involving xylazine held steady at 1.88 per 100,000 in 2023 and 1.89 in 2024—representing an increasing proportion of fentanyl-involved deaths over time.

## Discussion

The US overdose crisis is undergoing a historically significant transition. After experiencing steady increases for nearly half a century, our findings suggest that US overdose mortality finally dipped below the Jalal-Burke exponential growth curve, in the midst of sharp declines occurring in 2023 and 2024. All four waves of the overdose crisis are now in decline, with wave four— defined by deaths involving fentanyl with stimulants—beginning to reverse in 2024. However, deaths involving certain drugs, including methamphetamine and cocaine without fentanyl, and xylazine, continue to rise.

From the inception of the concept of the exponential growth curve, it was theorized to be an inherently limited phenomenon; exponential growth must eventually flatten out due to physical constraints (Jalal et al., 2018). Jalal and Burke also speculated that the observed exponential growth curve may actually represent the early portion of a logistic curve, which will emerge into an ‘S’ shape with time (Burke & Jalal, 2022).

Various mechanisms have been proposed to explain both previous exponential growth and more recent decreases in death rates. Exponential increases were likely related to increasingly early and large magnitude upticks in overdose mortality in each subsequent US birth cohort (Jalal et al., 2020) as well as waves of increasingly potent opioids in the illicit drug market (Ciccarone, 2021; J. Friedman & Shover, 2023). Recent decreases have been hypothesized to reflect changes in the number of remaining susceptible individuals, drops in fentanyl potency, increasing tolerance, shifting use practices (e.g. increased smoking rather than injecting of opioids) and health policies and harm reduction practices, such as increased access to naloxone and medications for opioid use disorder (J. R. Friedman et al., 2026; Stringfellow et al., 2025; Vangelov et al., 2026). The relative importance of each of these factors in explaining decreases remains to be determined, but the permanence of continued decreases will likely depend on future shifts across each of these areas. It also remains to be seen if these various mechanisms result in a bell-shaped curve wherein overdose mortality returns to much lower levels seen in prior decades, or if they level out in a pattern more characteristic of a logistic curve.

Although recent decreases are substantial, policymakers, clinicians, and public health officials should be prepared for the possibility of a return to exponential growth, especially if novel synthetic drugs once again change the risk environment of drug use. Novel classes of illicit substances have already seen pockets of increased prevalence in recent years, and have the potential capacity to drive increasing overdose death rates should they begin to proliferate widely in the illicit drug market. Many of the underlying social, economic, and political structural conditions that permitted the overdose crisis to grow exponentially for so long remain deep-seated in the US (Dasgupta et al., 2018; Caulkins, 2022; J. R. Friedman et al., 2024). Of note, deaths involving methamphetamine and/or cocaine (without fentanyl) and xylazine are still increasing, highlighting that the nature of the crisis continues to present new public health challenges requiring updated interventions. The rising importance of stimulants and xylazine may warrant increased focus on nonfatal, rather than fatal health harms.

Furthermore, despite progress, overdose death rates remain at about 400% of the magnitude seen in the year 2000 and continue to be far higher than those in other developed nations (Aziani & Caulkins, 2023; Gumas, 2025). The nearly 80,000 lives lost to overdose in 2024 remain a staggering quantity of preventable mortality. The sharp exponential growth seen in the US in recent decades amidst transition from agricultural to synthetic drug products could also be relevant to public health planning in other countries that may be experiencing initial increases in overdose deaths amidst the proliferation of nitazenes and other synthetic opioids(Giraudon et al., 2024).

## Limitations

Limitations of this study include the provisional nature of 2024 data, which may change slightly in final numbers. Caution should also be used in interpreting this dip below the curve to imply continued decreases; only time will tell if this represents the beginning of sustained decreases, or a temporary downward shock.

As is standard among analyses using national overdose death data, our codes for ‘fentanyl’ refer to synthetic opioids broadly, but are understood to represent majority fentanyl and fentanyl analog deaths(J. Friedman & Shover, 2023). Similarly, our ‘methamphetamine’ category refers to psychostimulants, known to be mostly methamphetamine(Han et al., 2021). Codes for ‘xylazine’ involved deaths may also include those corresponding to medetomidine and other related agents(J. R. Friedman, 2025). Data are lagged, so they do not include trends through the past year, and data on certain newly emergent drugs, such as medetomidine, are not yet available in a specific fashion.

## Conclusion

Our findings suggest that, rather than simply representing a return to the Jalal-Burke exponential growth curve, recent decreases in overdose deaths represent the first significant, large-magnitude deviation below exponential growth projections since the curve was defined using data beginning in 1979. These trends represent a very positive development. However, the rising share of deaths involving stimulants and xylazine highlights that challenges in the US drug crisis are shifting, nonfatal health harms from addiction may be rising in importance, and exponential growth could be seen again in the future if shifts in the illicit drug market—together with underlying social, economic, and political conditions—trigger renewed increases.

## Data Availability

All data used in this study are publicly available through the US National Vital Statistics System.

## Funding

JRF received funding from the National Institute on Drug Abuse (DA049644; U01DA063078) and the National Institute of Mental Health (MH101072). JP received support from the National Institute on Drug Abuse of the National Institutes of Health (R01DA057289 and U01DA051126). DC received funding from the National Institute on Drug Abuse (R01DA054190 and R21DA064011). CLS was supported by National Institute on Drug Abuse (R01DA057630; K01DA05771). AB was supported by National Institute on Drug Abuse (R33DA061260 and DP2DA049295). SAS received funding from the National Institute of Drug Abuse (U01DA063078; R33DA061260). Funders had no role in the design and conduct of the study; collection, management, analysis, and interpretation of the data; preparation, review, or approval of the manuscript; and decision to submit the manuscript for publication.

## Conflicts of Interest

JP and DC are associated editors at the International Journal of Drug Policy, were not involved in the peer review of this article, and had no access to information regarding its peer review. Full responsibility for the editorial process for this article was delegated to (an)other journal editor(s). JP reported receiving personal fees from the Washington-Baltimore High Intensity Drug Trafficking Areas program, Elsevier, Wiley, Rutgers University, Arizona State University, the University of Southern California, Queensland University, the National Network of Public Health Institutes, Alta Mira Recovery programs, and Dartmouth University, and nonfinancial support from NIH/NIDA, the University of Florida, Rx Summit, the American College of Neuropsychopharmacology, and the Reagan-Udall Foundation for the FDA during the conduct of the study. DC reports personal fees from Celero Systems and Motley-Rice LLC outside the submitted work. All other authors declare no conflict of interest.

## Notes

### Author Declarations

This study was deemed exempt from review and informed consent by the University of California, San Diego Institutional Review Board as it uses only publicly available, deidentified records.

### Summary of Updates

Lengthened discussion, methods, increased number of references. Updated title.

